# SARS-CoV-2 Vaccine-Induced Antibody Response and Reinfection in Persons with Past Natural Infection

**DOI:** 10.1101/2021.05.18.21257259

**Authors:** Nitu Chauhan, Ajeet Singh Chahar, Prem Singh, Neel Sarovar Bhavesh, Ravi Tandon, Rupesh Chaturvedi

## Abstract

Several studies have shown that subjects with a history of severe acute respiratory syndrome coronavirus 2 (SARS-CoV-2) infection had significantly higher antibody titers than previously uninfected vaccinees after vaccination with mRNA vaccine. Yet no information is available for other vaccines.

In the current observational cohort study, 105 health care workers who had received Covishield an Adeno associated viral vector-based DNA vaccine were enrolled at Sarojini Nadu Medical College Agra, India. The study included 40 (23 men and 17 women) subjects with a previous history of SARS-CoV-2 infection and 65 participants (39 men and 26 women) who were not infected previously. Both the groups received the adenovirus vector vaccine ChAdOx1-S recombinant vaccines (Covishield, Astra Zeneca). The levels of SARS-CoV-2-anti-spike-IgG-antibodies titer were measured using Electrochemiluminescence immunoassay on Roche platform as arbitrary units per milliliter (AU/ml).

After 28 days of the second dose, subjects with no previous SARSCoV-2 infection showed a significantly lower level of circulating anti-spike-IgG-antibody titers compared to previously infected participants. After the second dose, we also observed a significant increase in SARS-CoV-2 infection in subjects with no prior history of SARS-CoV-2 infection compared to subjects with a previous history of natural infection.

The most important observation of the study is a low percentage of infection in previously infected subjects. The finding of the study also indicates the presence of robust humoral memory response in previously infected patients.

## Introduction

Several studies have shown that subjects with a history of severe acute respiratory syndrome coronavirus 2 (SARS-CoV-2) infection had significantly higher antibody titers than previously uninfected vaccinees after vaccination with mRNA vaccine1–3. Yet no information is available for other vaccines. It is important to know if vaccinated subjects with a previous history of SARS-CoV-2 infection are better protected against SARS-CoV-2 infection compared to vaccinated individuals with no previous history of infection.

## Methods and Result

In the current observational cohort study, 105 health care workers who had received Covishield an Adeno associated viral vector-based DNA vaccine were enrolled at Sarojini Nadu Medical College Agra, India. The study included 40 (23 men and 17 women) subjects with the previous history of SARS-CoV-2 infection (30 symptomatic and 10 Asymptomatic subjects). The mean age of previously infected participants was 37 ± 2.5 years (95% confidence interval [CI], 31.6 to 42.3). Symptomatic or asymptomatic subjects were characterized on the basis of documented evidence of RT-PCR or Rapid antigen test and clinical symptoms or antibody titers at the time of vaccination (0 days). The current study also included 65 participants (39 men and 26 women) who were not infected previously. The mean age of uninfected participants was 42 ± 1.6 years (95% CI, 38.7 to 44.4).

Both the groups received the adenovirus vector vaccine ChAdOx1-S recombinant vaccines (Covishield, Astra Zeneca). Serum samples were obtained at the day of vaccination (0 days), 28 days after the first and second dose respectively. The levels of SARS-CoV-2-anti-spike-IgG-antibodies titer were measured using Electrochemiluminescence immunoassay on Roche platform as arbitrary units per milliliter (AU/ml) after.

After 28 days of the second dose, subjects with no previous SARSCoV-2 infection showed a significantly lower level of circulating anti-spike-IgG-antibody titers compared to previously infected participants (mean-2881 AU/ml; 95%CI, 2286-3475; mean-540; 95%CI, 318-763 respectively, p<0.0001; Figure 1A). All the subjects with previous natural infection showed a significant increase in anti-spike-IgG-antibodies titer level after the second dose compared to pre-vaccination anti-spike-IgG-antibodies titer (mean-3140 AU/ml; 95%CI, 2429-3851; mean-79 AU/ml; 95%CI, 33-124 respectively, p<0.0001; Figure 1B). We also observed that after the second dose, 22% (14/65) of previously uninfected subjects got infected (mean days for infection post-vaccination; 65 days) with mild symptom in the second wave of pandemic compared to 2.5% in the previously infected group (21/40; P= 0.0082; Figure 1C). Thirteen subjects out of 14 infected subjects did not show an increase in levels of titers after the second dose compared to the first dose (Figure 1D).

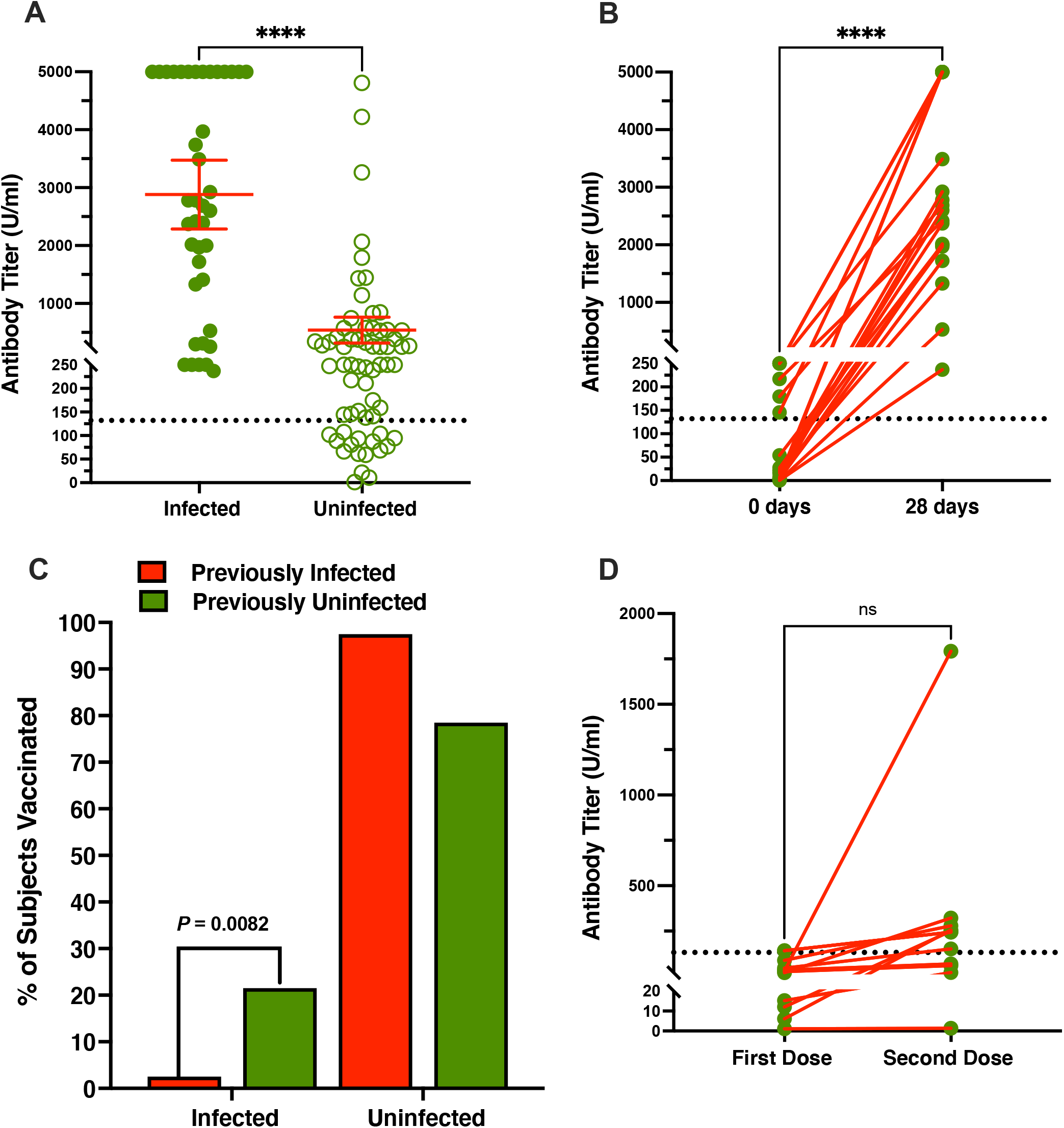
Titers of circulating severe acute respiratory syndrome coronavirus 2 (SARS-CoV-2) anti-spike IgG antibodies were measured after vaccination with Covishield: After 28 days of second dose of vaccine in subjects who were previously infected or uninfected (Panel A); in previously infected individuals at the day of first dose (0 day) and after 28 days of second dose (28 days) of vaccine (Panel B); Percent of subjects who got SARSCoV-2 infection after 28 days of second dose in both groups (Panel C). in serum samples from subjects collected from previously uninfected subjects who got infected after second dose of vaccine. Serum samples were used obtained after 28 days of First and Second dose (Panel D). Dotted line is a cut-off recommended by Federal drug and food administration for therapeutic antibody titer for plasma therapy (>132U/ml). Previously uninfected (*n = 65*), previously infected group (*n = 40*), previously infected asymptomatic (*n = 10*), and symptomatic (*n = 30*) subjects, post vaccination infected subjects; previously uninfected (*14 out of 65; Panel E*) and previously infected (*1 out of 40)*, **** (*p;* Two-tailed < 0.0001). Samples with titer level 250 were diluted to 1:20 for repeat measurement.

## Discussion

The most important observation of the study is the low percentage of infection in previously infected subjects. The finding of the study also indicates the presence of robust humoral memory response in previously infected patients. This study suggests a need for monitoring the levels of antibody titers in post-vaccinated previously uninfected subjects to achieve a desirable level of protection in a population. Subjects with lower levels of antibody titer may require a second booster to elevate the antibody levels.

## Data Availability

All Raw and analyzed data are available at the Department of Transfusion Medicine, S. N. Medical College, Agra in excel format.

## Notes

### Competing Interest Statement

The authors have declared no competing interest.

### Clinical Trial

This is part of ongoing government sponsored health worker immunization program.

### Funding Statement

No external funding is provided to the institution or authors for submitted work.

### Author Declarations

Institutional Ethics Committee, S. N. Medical College Agra, E. C. Registration No.: ECR/1409/Inst?UP/2020 Study Approval No.: SNMC/IEC/2021-09

## Reference

1. Wise J. Covid-19: people who have had infection might only need one dose of mRNA vaccine. BMJ 2021;372: 308-n308.

2. Krammer F, Srivastava K, Alshammary H, et al. Antibody responses in seropositive persons after a single dose of SARS-CoV-2 mRNA vaccine. N Engl J Med 2021; 384:1372–1374.

3. Anichini G, Terrosi C, Gandolfo C, et al. SARS-CoV-2 Antibody Response in Persons with Past Natural Infection. N Engl J Med 2021; Apr 14; NEJMc2103825

